# The roadmap for implementing value based healthcare in European university hospitals - consensus report and recommendations

**DOI:** 10.1101/2021.05.18.21257238

**Authors:** Yolima Cossio-Gil, Tanja Stamm, Maisa Omara, Carolina Watson, Joseph Casey, Alex Chakhunashvili, François Crémieux, María Gutiérrez-San Miguel, Pascal Kahlem, Samuel Keuchkerian, Valerie Kirchberger, Virginie Luce-Garnier, Dominik Michiels, Matteo Moro, Barbara Philipp-Jaschek, Simona Sancini, Jan Hazelzet

## Abstract

Value based healthcare (VBHC) aims at improving patient outcomes while optimizing the use of hospitals’ resources among medical personnel, administrations and support services through an evidence-based, collaborative approach.

In this paper, we present a blueprint for the implementation of VBHC in hospitals, based on our experience as members of the European University Hospital Alliance (EUHA). The EUHA is a consortium of nine large hospitals in Europe and aims at increasing quality and efficiency of care to ultimately drive better outcomes for patients. The blueprint describes how to prepare hospitals for VBHC implementation, analyses gaps, barriers and facilitators and explores the most effective ways to turn patient pathways to a process that results in high value care. Using a patient centric approach, we identified four core minimum components that must be established as cornerstones and seven organisational enablers to waive the barriers to implementation and ensure sustainability. The blueprint guides through pathway implementation and establishment of key performance indicators in six phases, which hospitals can tailor to their current status on their way to implement VBHC.

## Introduction and background

Measuring outcomes that matter to patients is one of the cornerstones of value-based healthcare (VBHC)^1^. VBHC puts outcomes at the center of the healthcare process and in relation to costs. It so determines the value of medical services which should be the driver for performance improvement in healthcare. Thus, VBHC contributes to the sustainability of healthcare systems and claims for reforming and re-constructing health systems globally^2^. Aligning healthcare approaches, focussing primarily on patient outcomes, can transcend quality, increase efficiency and enable a patient-centric approach while reducing costs and burden on already overstretched support services^1^.

Porter and Teisberg who pioneered VBHC argued that the transformation should be based on six interrelated elements: (i) organize into integrated practice units (IPUs), (ii) measure outcomes and costs for every patient, (iii) move to bundled payments for care cycles, (iv) integrate care delivery systems, (v) expand geographic reach and (vi) build and enable information technology platform^3,4^. A similar approach has been followed in the so-called Quadruple Aim Model^5^ that focuses on increasing population health, while reducing the cost per capita and improving the experience of patients and caregivers. However, while the VBHC elements provide a broad view of the systems’ parameters that need to be considered, implemention still remains largely in pilot phase^6,7^ and hospitals show diverse maturity levels. Thus, a general roadmap of transformational measures towards VBHC is lacking.

In 2017, nine leading European university hospitals established the European University Hospitals Alliance (EUHA), setting out a commitment towards excellence in healthcare, education and research, with the overall aim to improve value of care in European. One of EUHA’s working groups focuses specifically on value, with an emphasis on pathways and outcomes. It was formed to engage in defining the minimum requirements to achieve efficient implementation of VBHC in the environment of university hospitals, assuming that hospitals in the different countries would face similar ressources and barriers. Furthermore, our ambition as a working group was to serve the hospital communities in their efforts to ensure that a patient-centric approach is taken while setting minimum requirements to increased efficiency when implementing VBHC measures. Moreover, we aimed at supporting other hospitals to start, integrate and further develop VBHC within their institutions and healthcare systems. We also aimed to define the training needs regarding VBHC for different healthcare professionals. We therefore established a generic roadmap (“blueprint”) for the implementation of VBHC in a hospital, including the different phases and the identification of possible enablers and barriers. Since there is no single definition of VBHC or of the meaning of value in a health context, we used a definition from the European Commission experts^8^. We also considered, that no matter how exact the definition of value was, our proposed blueprint would be of use for organizations in moving towards an outcome-driven patient-centered system.

## Methods

We performed an international, multicenter consensus process. The multidisciplinary working group consisted of two convenors (YC, JH), a methodologist (TS) and experts in patient-reported health outcomes, care process improvement and care pathway design. Experts’ backgrounds spanned from nurses, medical doctors, process engineers, statisticians, hospital managers and outcomes researchers working in one of the nine EUHA university hospitals. An average of two experts participated per hospital.

The working group members were asked to indicate whether any step was considered, planned and/or implemented within their institution. We then met seven times face-to face or virtually, added field-visits, where possible, separated by periods of two months. During the face-to-face meetings at the different hospitals, we analysed the level of implementation of VBHC, exchanged knowledge on real evidence and learned from each other. Within this iterative process, we selected specific critical components for the implementation of VBHC in our hospitals. We invited external experts to discuss the following specific topics to our meetings: team collaboration, service design, outcomes measurement, lean methodology and organizational transformation, process improvement, VBHC strategy and tactics, and information technology. All experts’ contributions acted on a non-commercial basis. We then drafted a process for implementing VBHC. In email rounds, we asked the working group members to comment on the draft version of the blueprint document until we reached the final version. In case of contradictory comments, the convenors and the methodologist discussed the pros and cons of each argument until consensus was achieved.

## Results

We identified eight mandatory components to implement VBHC in a hospital (Figure 1) and grouped them into four main areas: the first three areas refer to Porter’s six core elements^9^ and a forth one was added by us. The first main area refers to **(i) organizing care pathways**. This is related to the IPUs recommended by Porter. Implementing a transformation to IPUs can become expensive and time consuming in university hospitals. It also requires extensive organizational and cultural changes in the way care is delivered. Therefore, we recommend starting with redirecting the process of attention in the form of a continued assistance by clinical condition. Mutual visits/exchange of staff with participation in care in a previous or subsequent unit can enhance the IPU mindset. However, in this document, we consider pathways rather than IPU. The second area is **(ii) collecting a set of outcomes**, including clinical outocmes, patient-reported outcomes measures (PROMs) as well as experience measures (PREMs), process indicators and in a later stage also costs at the patient level. **(iii) Building an Information platform** is the third area. We recommend to enable the collection of PROMs integrated within the patient pathway and the visualisation of these data using dashboards where indicators are represented. This information platform must allow to communicate and provide feedback regarding PROMs to clinical teams and also feedback to patients about their own health status. The set up of a datawarehouse enviroment on VBHC, taking into account that process indicators and general care information (EMR) need to be linked to CROMs and PROMs. Another forth area is **(iv) actively using short-term and long-term outcomes for clinical decisions and for improving care, with the aim for a patient-centric approach**. Cultural change towards actively using the PROMs and PREMs with the patients, and enabling shared decision-making tools is mandatory in order to grasp significant patient-centered care.

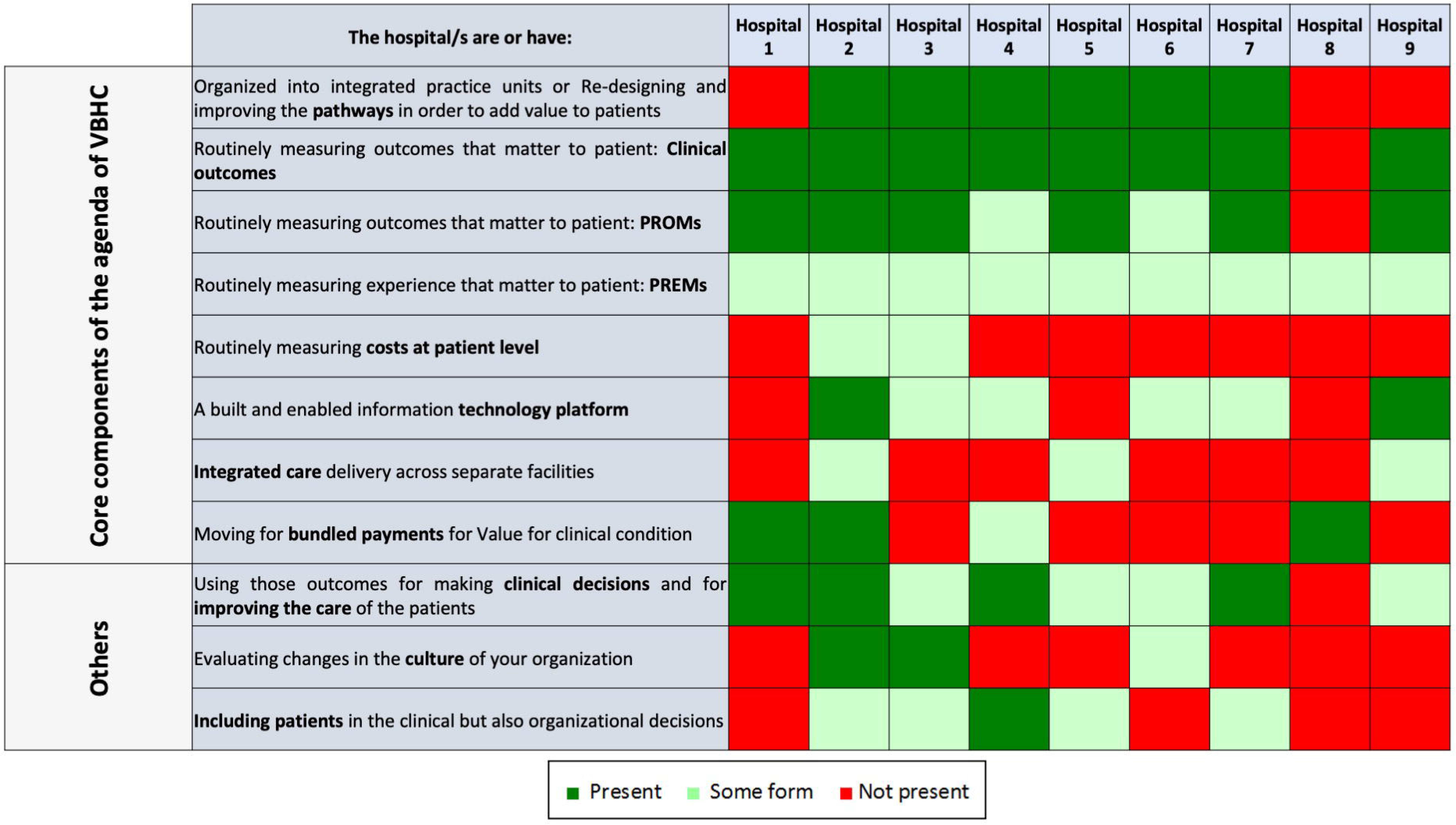

Based on these four main areas, we structured the implementation in six development phases (Figure 2). While phase 1 corresponds to the preparation of the whole organization for VBHC, the following five phases entail the concrete implementation of the clinical pathway.

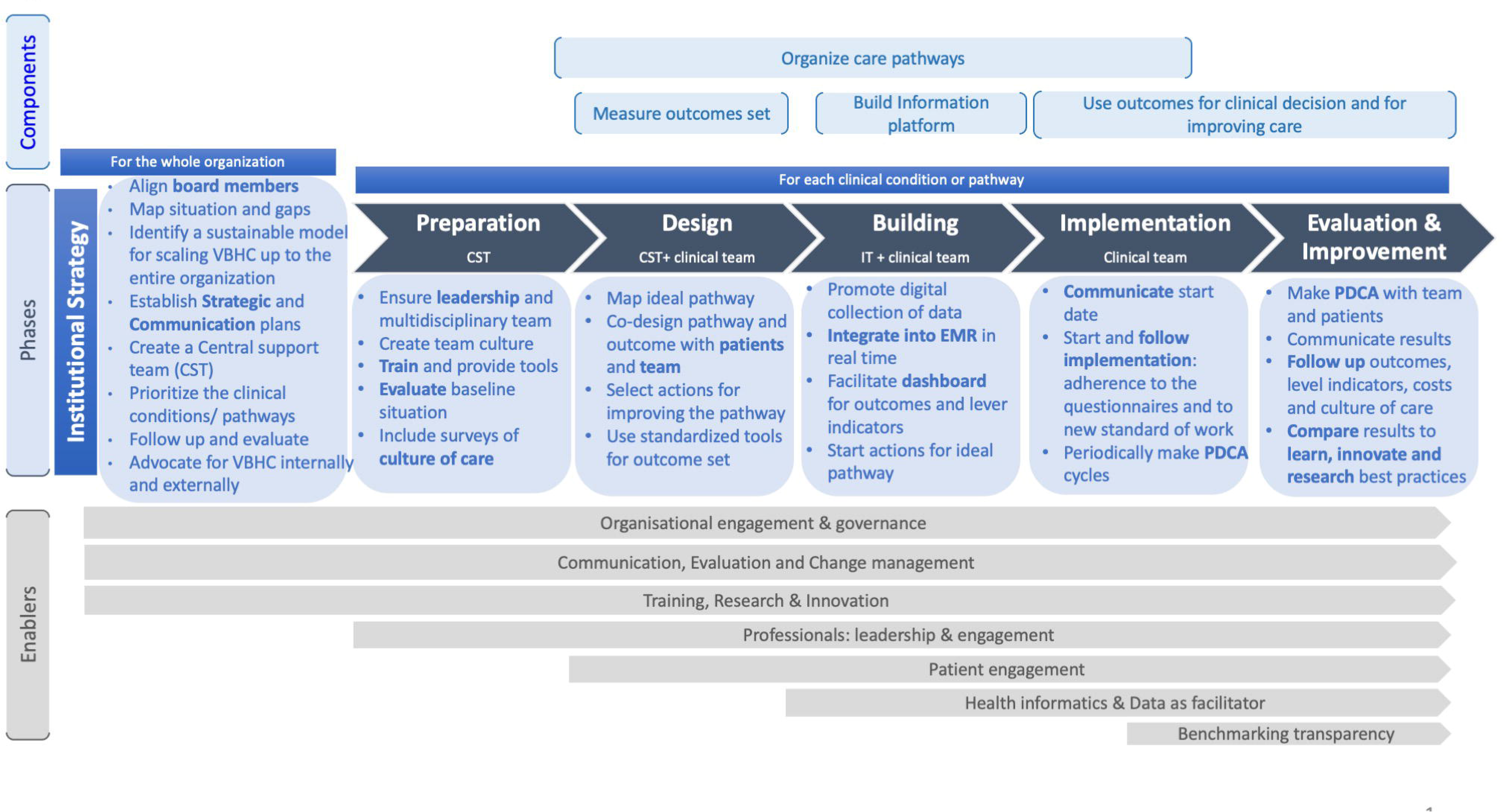

### Phase 1: Preparation of the whole organization for VBHC: Institutional Strategy

In this phase, the organization sets up the strategic plan for the implementation of VBHC, including evaluation and follow-up of the maturity and readiness to transform towards VBHC. The main actors in this phase are the board members, who create a strategic umbrella for implementing VBHC in all levels of the organization. If the organization is not yet ready for the substantial change, we recommend starting with pilots. Also, we suggest to align board members, map the current situation and analyse the gaps using an evaluation tool of the maturity of the organization in relation to value, e.g. the Value Accelerators. In addition, it is important to identify a sustainable model for scaling VBHC up to the entire organization and consider starting value purchasing methods and negotiating the payment for value. Parallel to this process, the hospital should set up a communication strategy (internal for hospital staff and external for the population, for the insurance agencies, government and other providers) and advocate for the VBHC inside and outside of the hospital.

A Central Support Team (CST) should be created with at least one strategic and one operation lead. They should be trained to be able to help all the clinical leaders with the implementation of each clinical pathway. Staffing these teams is essential. The team size can vary according to the size of the organization. The multidisciplinarity of the suport team is recommendable as clinical point of views are essencial in order to connect with the clinical leads and enhance the cultural change but may be “too deep” into the hospital culture. New mindsets from other professional profiles may be of added value in order to bring on fresh inputs in the re-organization of the care pathways that come with VBHC.

Hospitals need to prioritize the clinical conditions/pathways. Board members should decide which one to start with, in order to apply the appropriate changes at the organisational level. The best strategy in each specific case should be defined (to go in deep with the pathway or identifying quick wins). We recommend to start with the clinical pathway where clinical leads show a great commitment and the team is engaged. Once the organization is gaining experience in the implementation, other criteria can be added, if there is room for improvement (e.g. fragmented care, lack of standardization in the procedures, results, costs, etc). Finally in this phase, we need to set up a follow up and evaluate the changes in the organization towards value.

### Phase 2: Preparation of each clinical pathway

This phase includes the steps to prepare the team for starting a new working model and for measuring the baseline situation of the costs, team culture and patients experience prior to implementing VBHC. The main actors in this phase are the members of the central support team. We recommend to ensure leadership and multidisciplinary participation, including physicians, non-physician health professionals including nurses, porters, administrative staff, social care, business intelligence/data managers, pharmacy. Hospitals should choose leader/s and some should sign an agreement of commitment between the clinical lead and the organization. Goals and expectation of the team members must be assessed and potential barrieres identified. Team culture should faciliate communication between all the people involved in the clinical pathway and using comunication tools should ensure that everyone receives the information. The ideas that arise from people that are not part of the improvement/working group are also included. It is important to ensure their engagement during the process and in decision-making.

Hostpitals should provide training and tools on methodologies to improve quality and processes, including, but not limited to, shared decision-making, culture of continuous improvement, communication with the patient and other tools that help the team to lead the changes. Furthermore, patients should be clustered into rational groups. Organizing care into pathways requires the ability to assign patients to pathways. Sometimes this is straightforward but not always. We recommend to group them according to a defined logical model (for example, using SNOMED codes or other grouping parameters).

The hospital should evaluate the baseline situation, including the outcomes which are measured, and conduct surveys on the current state of the culture of care. VBHC is about improving patient treatment results and costs, but also reducing the burden on professionals and improving satisfaction with their work. Therefore, in addition to measuring the baseline costs, PREMs (Patient Reported Experience Measures) and the process indicators of each clinical pathway, we recommend to measure the clinical team culture and work environment.

### Phase 3: Design (by the CST and the clinical team)

In this phase, the outcome set and the standard of care for patients with the selected clinical condition should be defined. It should include process indicators to measure over (lead times, reports, presence of key interventions, among others) and the underuse of health care, detecting the possible root cause of outcome deficiencies and identifying the appropriateness of clinical practice linked to the outcomes. Surrogate measures, understood as those that are related/associated to higher level measures, should be taken into acount. For instance, in the case of diabetes, HbA1c can be an example of such a surrogate measure since it has a profound impact on the higher-level measures such as mortality and morbidity (e.g. unwanted complications as a result of an intervention due to other clinical condition). In this phase, the central support team should help the clinical team to find a better standard of work and to decide the outcomes of value that will be measured and monitored. Important is to co-design the pathway and outcomes with patients and team by considering what matters to patients. Focus groups, journey map, surveys to patient, literature reviewing could be some of the methodologies that could help in this stage. A countinuous improvement process should be stated based on the outcomes measured.

### Phase 4: Building (by the IT and the clinical team)

In this phase, the IT and business intelligence staff create the solutions for collecting, analysing and visualizing the outcomes and the process indicators. In parallel, the clinical team should start to implement the main changes to improve the pathway or circuit of attention. Note that the improvement of the pathway also continues in all the other phases in an iterative manner.

Outcome data need to be integrated in the electronic health records. IT experts need to prepare EMRs to check key interventions. We recommend to move from retrospective to prospective indicators including costs for an individual patient stay/journey. Dashboards or other tools could help the team to easy visualize indicators of processes and outcomes to make the continuous improvement. Visualizing outcomes will help clinicians to improve communication with patients as well as monitoring in between clinical visits. Patients should have access to their own outcomes and evolution over time.

### Phase 5: Implementing (by the clinical teams)

Implementing VBHC on an organizational level as well as in a pilot environment requires a multi-level effort. In this phase, the team should focus on getting the capture of PROs as well as the implementation of changes to the health care pathway. It should also already at this point in time consider the continuous monitoring of both. The main actors here are the clinical lead and the clinical team. Measuring outcomes and fostering discussions on treatment pathways will facilitate the culture and organizational changes that were planned in phases 2 and 3 (mainly organizing pathways, breaking profile-based culture, patient-focused culture).

Continuous improvement could be ensured through systematic “Plan Do Check Act” (PDCA) cycles: Follow the implementation process with indicators related to PRO measurement (i.e. compliance of the questionnaires) and to clinical appropriateness along the pathway (optimal timings, overuse and underuse). Regultar meetings where PDCA could be used as a tool and mindset for improvement, distinguishing PDCA cycles for data collection (i.e. involvement of patients and clinicians) and PDCA cycles for the pathway itself.

### Phase 6: Evaluation and improvement

In this phase, we recommend to use the outcomes and process indicators to evaluate the changes and follow up the improvement. Annually or biannually, PREMs, culture within the disease teams as well as costs should be assessed and compared with the baseline data. Patient feedback should be used to facilitate continuous improvement. Periodic meetings with the full team (every 2-3 months) to assess the aggregate outcomes and plan actions for improving them. We recommend to use a methodology to find the real cause of the problems and to prioritize the actions. Results should be communicated with teams, the board members, the rest of the organization and to external stakeholders according to the strategy defined earlier.

Results could be also compared to learn and innovate. Best practice examples could be identified and used to innovate research and training.

### Challenges and enablers

Table 1 summarizes the key gaps and barriers identified by the working group. We also propose mitigation strategies and recommendations on how to overcome each of them.

**Table 1.**
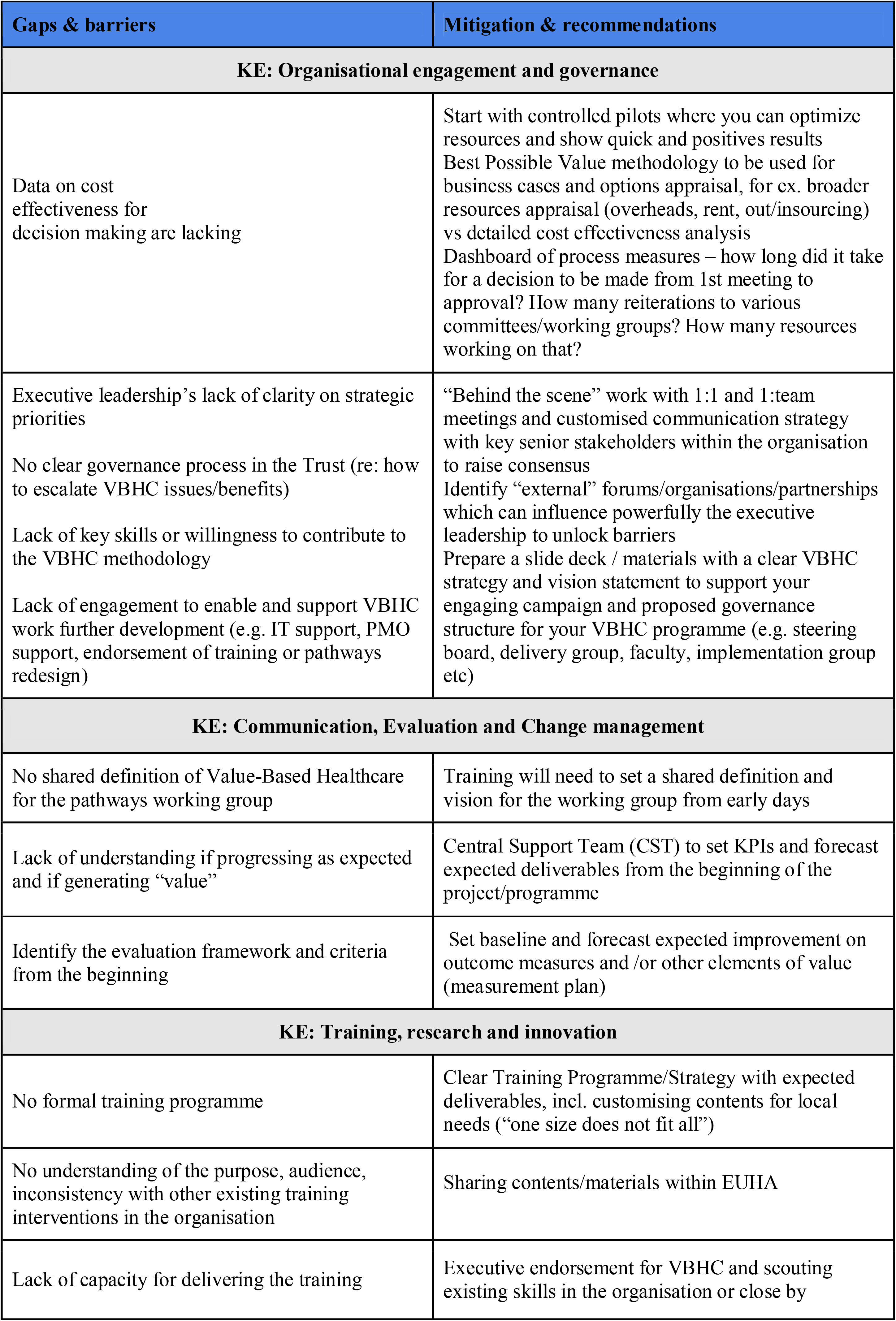
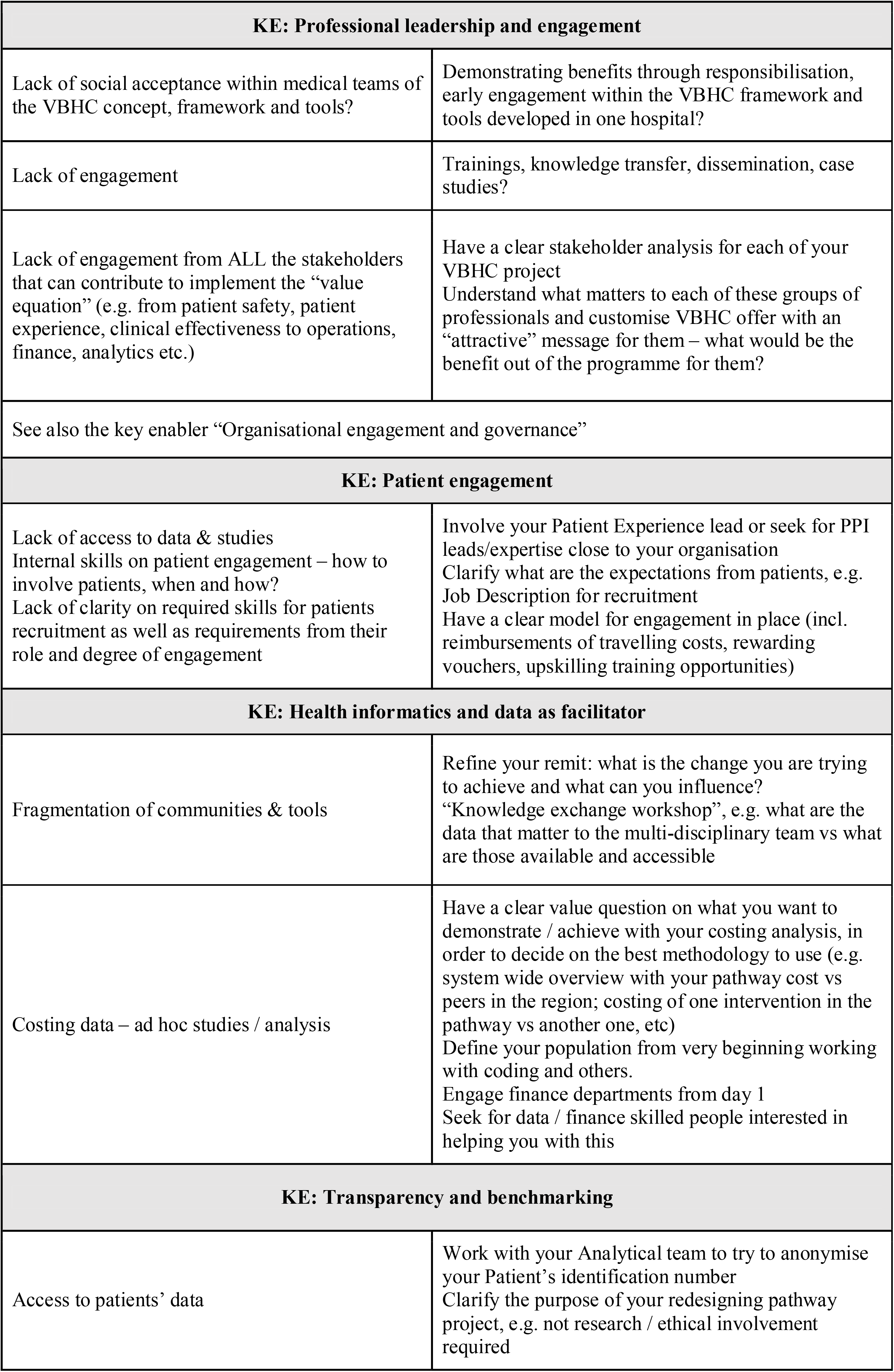
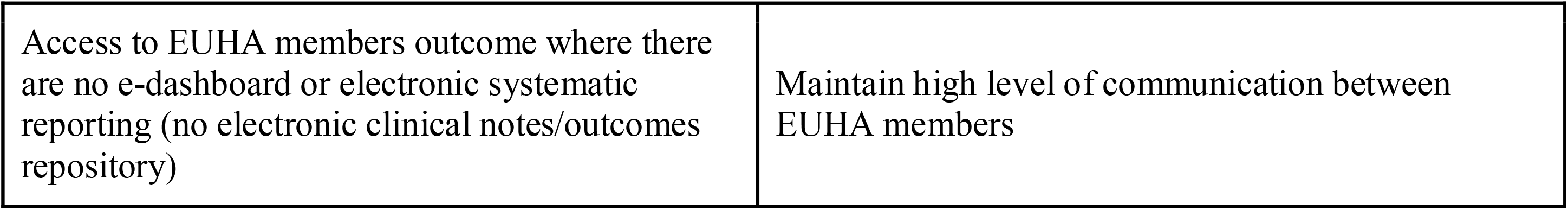
Gaps and barriers for each of the key enablers (KE) identified by EUHA Outcome & Pathways working group and mitigation strategies & recommendations to each of them.

## Discussion

This paper summarizes the development and implementation of a VBHC blueprint based on the consensus from nine of the largest university hospitals in the EU. This was driven by the opinions and experiences of the working group, and could be used as a strategic tool and guidance to university hospitals.

While prior studies have identified the importance of implementing VBHC, very little has been made in terms of guidelines to health care organizations and hospitals as to how implementation should be managed and sustained. Our roadmap provides for the first time, practical and applicable recommendations to each phase of the implementation of VBHC from a hospital management perspective. With this we added, in our opinion, an important component to Porter’s agenda.

This paper focuses on three main points of Porter’s agenda (IPU-outcomes-platform), but it addresses neither the cost-per-patient measurement, the organisation transformation towards a payment model based on value-based results/value based pricing, nor the integration and coordination with other services such as primary care, social care, and others. While we address the hospital perspective only, the implementation of VBHC needs to be embeded in a larger healthcare ecosystem. Implementing VBHC involves a broader range of elements than those described in this report (e.g. payments, etc). For all these reasons, this paper should be considered as an initial set of recommendations to foster building the bases at the hospital level, and future studies on the current topic are therefore recommended.

While top-down implementation experiences have shown poor response and adherence from patients, due to lack of commitment to improvement by the lower-level employees^10^ bottom-up experiences are usually pilots with difficulties in terms of consolidation and entrenchment of a general model or blueprint. From our point of view, hospitals must promote an implementation integrated in the practical clinical practice. The implementation needs the commitment and action of many actors of the healthcare system, beyond the hospital itself. Clinical teams and patients must have access to the data for making changes at the point of care and discuss the decision with patients. The hospital cannot change the payment system itself, but can either test different models to convince the payer or get prepared to face the model of the change when it comes.

It is important to understand that implementing VBHC should be based on an iterative process including evidence and a continious self-learning process in order to achieve the maximum patient-relevant medical benefits (outcomes) and minimize the costs. Accordingly, it could be more practical to start the process through a well-designed pilot in order to evaluate risks and opportunities on real life circumstances. It is recommended to embed the pilot as much as possible in generic systems of the whole hospital and let them grow/mature together, as scaling up the VBHC journey is completely dependent on this balance. Starting the pilot in a selected health condition or pathway where better circumstances are available (e.g. motivated clinical lead and engaged team) could be the way to intitiate a proper understanding for the process and how it can be implemented, although it might not reveal at the begining all chalenges which might be encountered. Moreover, the choice to start with selecting specific health conditions/care pathways or to have a complete VBHC transformation should be tuned to the specific situation of each instiution. Although adopting specific conditions and care pathway strategies then scaling up to other conditions seems to be a less risky approach, both models should be further studied and explored in the future including their related outcomes and costs.

Identifing a sustainable model for VBHC is a very important approach to visualize the future of VBHC within the organization and ensure the sucess of the system on the short and long term. Therefore, advocating for VBHC with providers and payers and setting a long-term plan with all stalkholders is an essential step.

## Data Availability

All data referred to in the manuscript and note links are available upon request.

## Acknowledgements

We acknowledge and are grateful for the valuable contribution of external organization experts:

Michele Van Der Kemp. Expert on Value Based Health Care Strategy and Tactics: Implementation

Muir Gray. Professor and Executive Director at Oxford Centre for Triple Value Healthcare

Gregory Katz. Chaired professor of Innovation & Value in Health at University of Paris School of Medicine, and President of the Consortium VBHC France

Ingeborg Griffioen. Industrial designer at Panton Medical design Agency - Patient Journey - service design

Mona Krichen. Director of the Organizational transformation engineering department. Agence Nationale d’Appui À la Performance (ANAP)

Stéphanie Aftimos. Lean Black Belt - Agence Nationale d’Appui À la Performance (ANAP) Tim Wilson. Managing Director at Oxford Centre for Triple Value Healthcare

Mathias Ekman. Director Industry Solutions Executive for Healthcare at Microsoft Western Europe.

Kris Vanhaecht. Associate Professor Quality in Healthcare at KU Leuven

